# Seasonal Trends of COVID-19 Deaths in Italy: A Confirmatory Linear Regression Study with Time Series Data from 2024/2025

**DOI:** 10.1101/2025.05.30.25328619

**Authors:** Marco Roccetti

## Abstract

This study builds on and extends a previous research conducted with the time series data of COVID-19 deaths in Italy in the period 2021-2024. In those earlier works, weekly COVID-19 deaths showed pronounced seasonal increasing variations of COVID-19 mortality, rising in summer and high fall to peak in early winter, and then declining in the late winter and spring periods. Using a linear regression model, this current study has validated the seasonal mortality pattern mentioned before. We present original findings achieved with recent data from 53 weeks starting with the end of May 2024 until the end of May 2025. They confirm a recurring seasonal pattern of COVID-19 deaths, rising in early summer and peaking in high fall. Then, we document a steady decrease, starting in mid-winter until the end of the spring period. Since there is little published work from Italy in this context, having clearly shown the seasons with common increasing variations of COVID-19 mortality offers a contribution towards helping that subset of *at risk* population who are seasonally vulnerable. Our present study indicates that the COVID-19 mortality trend observed in previous years is still continuing, influenced by several factors that need further investigation.

## 1. Introduction

On May 5 2023, the World Health Organization (WHO) announced the beginning of the COVID-19 post-pandemic era [1]. Nonetheless, at least in all the Western countries, with each new year the question arises again as to which season is the most prone to this specific disease. Usually in the winter, other respiratory viruses illnesses, like cold and flu, are common, tending to peak during this time. The main factors to which this is attributable are: i) colder temperatures that can weaken the immune system thus making people more susceptible to infections, and ii) an increased amount of individuals spending time indoors in closer proximity.

The same kind of question, about the possibility of the COVID-19 illness to follow a similar one-year seasonal pattern, rising and peaking in winter, has been subject of an intense scientific debate, with the emergence of two opposing ideas. On one side, some are in favour of the existence of a seasonal infection pattern focused only on the winter months [2], on the other side, many have brought to the attention of the scientific community evidences of several repeating outbreaks occurring in different seasons, even if they are more common and apt to be worse in winter [3]. We have had this discussion repeated many times over these years also in Italy, with the two sides proposing almost the same arguments mentioned above.

Nevertheless, in some recent works, conducted in Italy with the time series data of weekly COVID-19 deaths of the period 2021-2024, when the Omicron and post-Omicron variants were predominant, we have documented, instead, pronounced seasonal increasing variations of COVID-19 mortality rising in summer and high fall to peak in early winter, and then steadily declining in mid- and post-winter periods [4].

In this present study, based on the use of a linear regression model, we have validated statistically the seasonal COVID-19 mortality pattern mentioned before. In fact, with recent data from 53 weeks, starting with the end of May 2024 until the end of May 2025, we have shown an increasing trend of weekly COVID-19 deaths, rising in early summer and peaking in high fall/early wither, as well as noting a steadily decreasing downfall of the number of COVID-19 deaths beginning in mid-winter until the end of the spring.

Since there is little published work from Italy with these results, having shown the seasons with increasing variations of COVID-19 mortality offers a help to that subset of *at risk* population who are seasonally vulnerable. In essence, our linear regression study has clearly indicated that the COVID-19 mortality trend observed in those years when the Omicron and post Omicron sub lineages were predominant (i.e., 2021-2024) is still continuing, influenced by several factors (including: new variants, decreasing immunity from previous infections and past vaccinations, environmental conditions, dense people gathering, relaxation of public health measures, holidays, …) that need further investigation [5, 6].

The remainder of this paper is organized as follows. In the Data and Methods Section, we describe where our data come from, as well as the methodology we have used to analyze them. In the Results Section, we present the results we have obtained, while in the Conclusion Section we terminate this paper also discussing the limitations of our approach.

## 2. Data and Methods

In the following, we provide sufficient details on used data and methods to allow readers to replicate our results.

### 2.1. Data

We have decided to work with the time series data of the Italian COVID-19 weekly deaths which are easily downloadable from the repository maintained by the Italian Ministry of Health, at the following URL: https://www.salute.gov.it/new/it/tema/covid-19/report-settimanali-covid-19/. All the data are available on a per week basis.

Since in our earlier works we studied the previous period 2021-2024 [4], the aim of this research was to validate statistically the results of those previous works, by documenting the COVID-19 seasonal mortality variations and the deaths time trends, observable in the most recent 53 weeks from 16 May 2024 to 21 May 2025.

To permit to readers to have a comprehensive and unique view of all the used data, we report all this data in Table 1, indicating, respectively: the number of consecutive weeks, any given week, and the corresponding number of weekly COVID-19 deaths. The timeframe of interest (i.e., 16 May 2024 - 21 May 2025) was chosen to include the rise of a COVID-19 mortality increasing trend, its subsequent peak, as well as the final downward decline, as clearly shown in Fig. 1, where a visual representation of the same data of Table 1 is also offered.

**Table 1.** COVID-19 deaths time series data (Italy). From 16 May 2024 to 21 May 2025: totaling 53 weeks.

| # Week | Week: beginning date - end date | Deaths per week |
| --- | --- | --- |
| 1 | 16 May 2024 - 22 May 2024 | 8 |
| 2 | 23 May 2024 - 29 May 2024 | 10 |
| 3 | 30 May 2024 - 5 June 2024 | 10 |
| 4 | 6 June 2024 - 12 June 2024 | 17 |
| 5 | 13 June 2024 - 19 June 2024 | 14 |
| 6 | 20 June 2024 - 26 June 2024 | 21 |
| 7 | 27 June 2024 - 13 July 2024 | 18 |
| 8 | 4 July 2024 - 10 July 2024 | 33 |
| 9 | 11 July 2024 - 17 July 2024 | 40 |
| 10 | 18 July 2024 - 24 July 2024 | 53 |
| 11 | 25 July 2024 - 31 July 2024 | 54 |
| 12 | 1 August 2024 - 7 August 2024 | 87 |
| 13 | 8 August 2024 - 14 August 2024 | 100 |
| 14 | 15 August 2024 - 21 August 2024 | 99 |
| 15 | 22 August 2024 - 28 August 2024 | 135 |
| 16 | 29 August 2024 - 4 September 2024 | 75 |
| 17 | 5 September 2024 - 11 September 2024 | 97 |
| 18 | 12 September 2024 - 18 September 2024 | 93 |
| 19 | 19 September 2024 - 25 September 2024 | 112 |
| 20 | 26 September 2024 - 2 October 2024 | 85 |
| 21 | 3 October 2024 - 9 October 2024 | 100 |
| 22 | 10 October 2024 - 16 October 2024 | 117 |
| 23 | 17 October 2024 - 23 October 2024 | 116 |
| 24 | 24 October 2024 - 30 October 2024 | 108 |
| 25 | 31 October 2024 - 6 November 2024 | 96 |
| 26 | 7 November 2024 - 13 November 2024 | 86 |
| 27 | 14 November 2024 - 20 November 2024 | 61 |
| 28 | 21 November 2024 - 27 November 2024 | 47 |
| 29 | 28 November 2024 - 4 December 2024 | 46 |
| 30 | 5 December 2024 - 11 December 2024 | 44 |
| 31 | 12 December 2024 - 18 December 2024 | 43 |
| 32 | 19 December 2024 - 25 December 2024 | 29 |
| 33 | 26 December 2024 - 1 January 2025 | 31 |
| 34 | 2 January 2025 - 8 January 2025 | 45 |
| 35 | 9 January 2025 - 15 January 2025 | 44 |
| 36 | 16 January 2025 - 22 January 2025 | 58 |
| 37 | 23 January 2025 - 29 January 2025 | 43 |
| 38 | 30 January 2025 - 5 February 2025 | 24 |
| 30 | 6 February 2025 - 12 February 2025 | 25 |
| 40 | 13 February 2025 - 19 February 2025 | 29 |
| 41 | 20 February 2025 - 26 February 2025 | 13 |
| 42 | 27 February 2025 - 5 March 2025 | 17 |
| 43 | 6 March 2025 - 12 March 2025 | 25 |
| 44 | 13 March 2025 - 19 March 2025 | 16 |
| 45 | 20 March 2025 - 26 March 2025 | 17 |
| 46 | 27 March 2025 - 2 April 2025 | 20 |
| 47 | 3 April 2025 - 9 April 2025 | 6 |
| 48 | 10 April 2025 - 16 April 2025 | 8 |
| 49 | 17 April 2025 - 23 April 2025 | 9 |
| 50 | 24 April 2025 - 30 April 2025 | 1 |
| 51 | 1 May 2025 - 7 May 2025 | 13 |
| 52 | 8 May 2025 - 14 May 2025 | 13 |
| 53 | 15 May 2025 - 21 May 2025 | 5 |

**Fig. 1.**
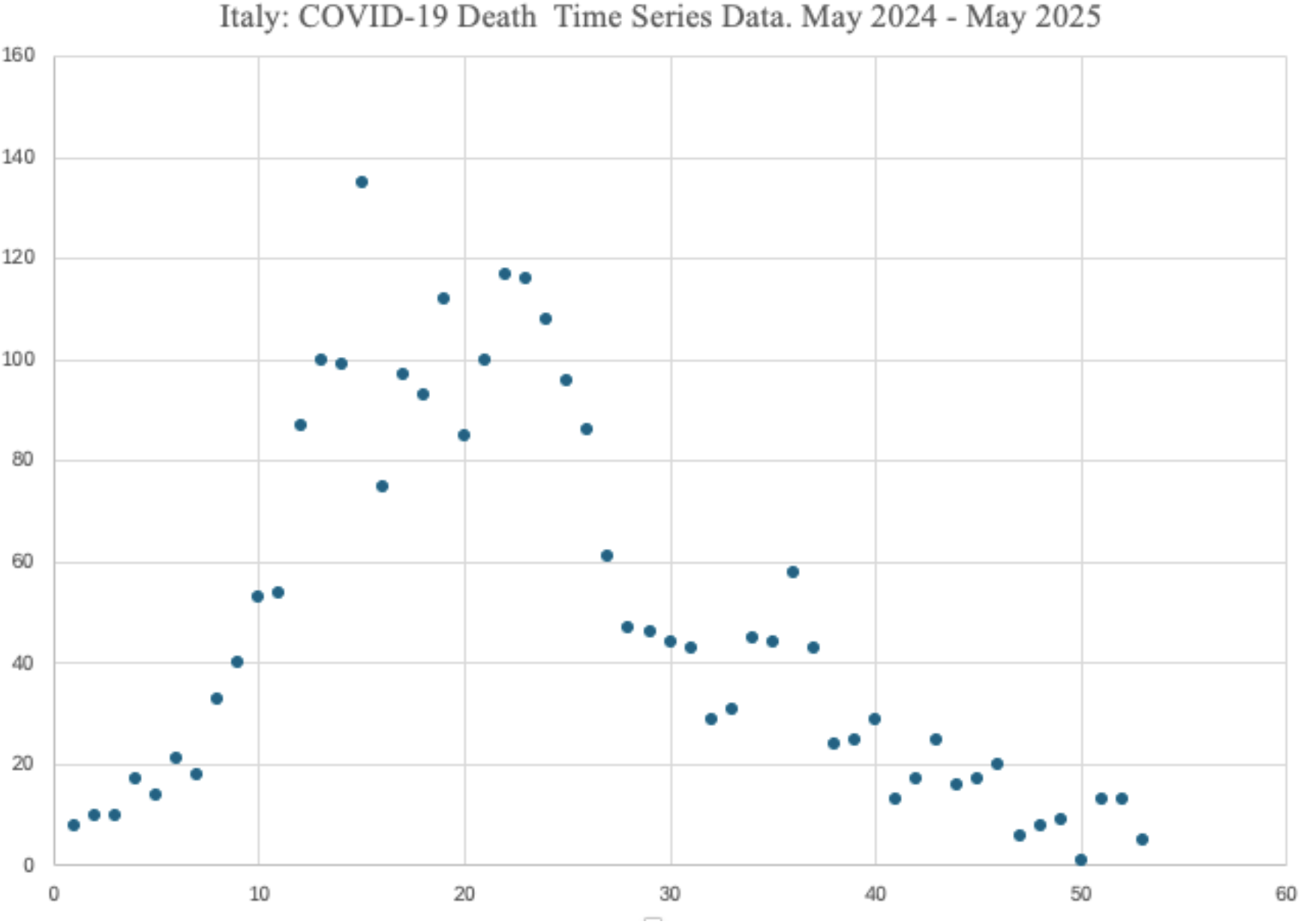
Visualization of the COVID-19 deaths data time series (Italy): 16 May 2024 - 21 May 2025. Y axis: number of COVID-19 deaths per week. X axis: weeks from 1 to 53.

### 2.2. Methods of analysis

With the data above, we fitted a piecewise linear regression model [7, 8], where the dependent variable was the number of confirmed weekly COVID-19 deaths and the independent variable was the number of weeks from the beginning to the end of the period of interest. In particular, we divided our 53 weeks into two separate periods. The first period extends from the first week under investigation (beginning on 16 May 2024) until the 27^th^ week (ending on 20 November 2024). The second period begins with the 28^th^ week (starting on 21 November 2024) and terminates with the 53^rd^ week (21 May 2025). In essence, the beginning of the first period was chosen in correspondence with the start of the rise of the increasing COVID-19 mortality trend, while the corresponding end point was the time when that increasing trend peaked. Similarly, the starting point of the second period was chosen in correspondence with the weeks immediately subsequent to the aforementioned mortality peak, while the end point corresponded to the time when the downward COVID-19 mortality trend had returned to baseline values.

Form a mathematical viewpoint, the piecewise, or segmented, regression model used to fit the COVID-19 deaths data of Fig. 1 follows the well-known formula:

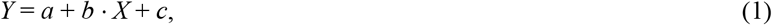

where *Y* corresponds to the number of the weekly COVID-19 deaths and *X* represents the passage of time measured in weeks. It is worth noting that *b* is the slope coefficient of the regression segment and indicates the steepness of that segment. It reflects the velocity with which a segment ascends (or descends) a mortality trend, informing about the change of the number of deaths for an increase of one week.

In this context, it is also important the role played by the coefficient of determination *r*^*2*^, which is needed to evaluate the goodness-of-fit of the simulated *Y* values (i.e., the entire segment) versus the measured *Y* values (i.e., the available deaths data of Fig. 1).

Finally, it is also worth noticing that we computed our regression function based on two separate segments (corresponding to the two different temporal periods mentioned above). This is due to an analysis of Fig. 1, from which it is evident that the COVID-19 deaths time series data follows two different trends: respectively, an ascending one in the first period and a descending one in the second period.

Summarizing, in the end, our model will return the values of the *b* parameter for both segments, which will be used, in turn, to evaluate the variation of the steepness of the COVID-19 death trends for the two seasonal periods under observation. With the values of *r*^*2*^ we will evaluate, instead, how well our segments fit the available data.

### 2.3. Informed consent statement

This study uses publicly available, aggregated data that contains no private information. Neither humans nor animals nor personal data are involved in this study. Therefore, ethical approval is not required.

### 2.4. Data availability statement

All the COVID-19 deaths data are downloadable from the repository of the Italian Ministry of Health: https://www.salute.gov.it/new/it/tema/covid-19/report-settimanali-covid-19/. Moreover, to allow readers to have an immediate access to all data, they have been also entirely reported in Table 1. All the results of this study are reproducible by using the linear regression method described in this Section. Further requests can be also addressed to the author.

### 2.5. Use of AI tools declaration

The author declares that he has not used artificial intelligence (AI) tools in the creation of this article.

## 3. Results

Fig. 2 and 3 show the regression segments obtained with our model for the two time periods of Fig. 1. Each Figure reports the dates of the beginning and of the end of the considered periods.

**Fig. 2.**
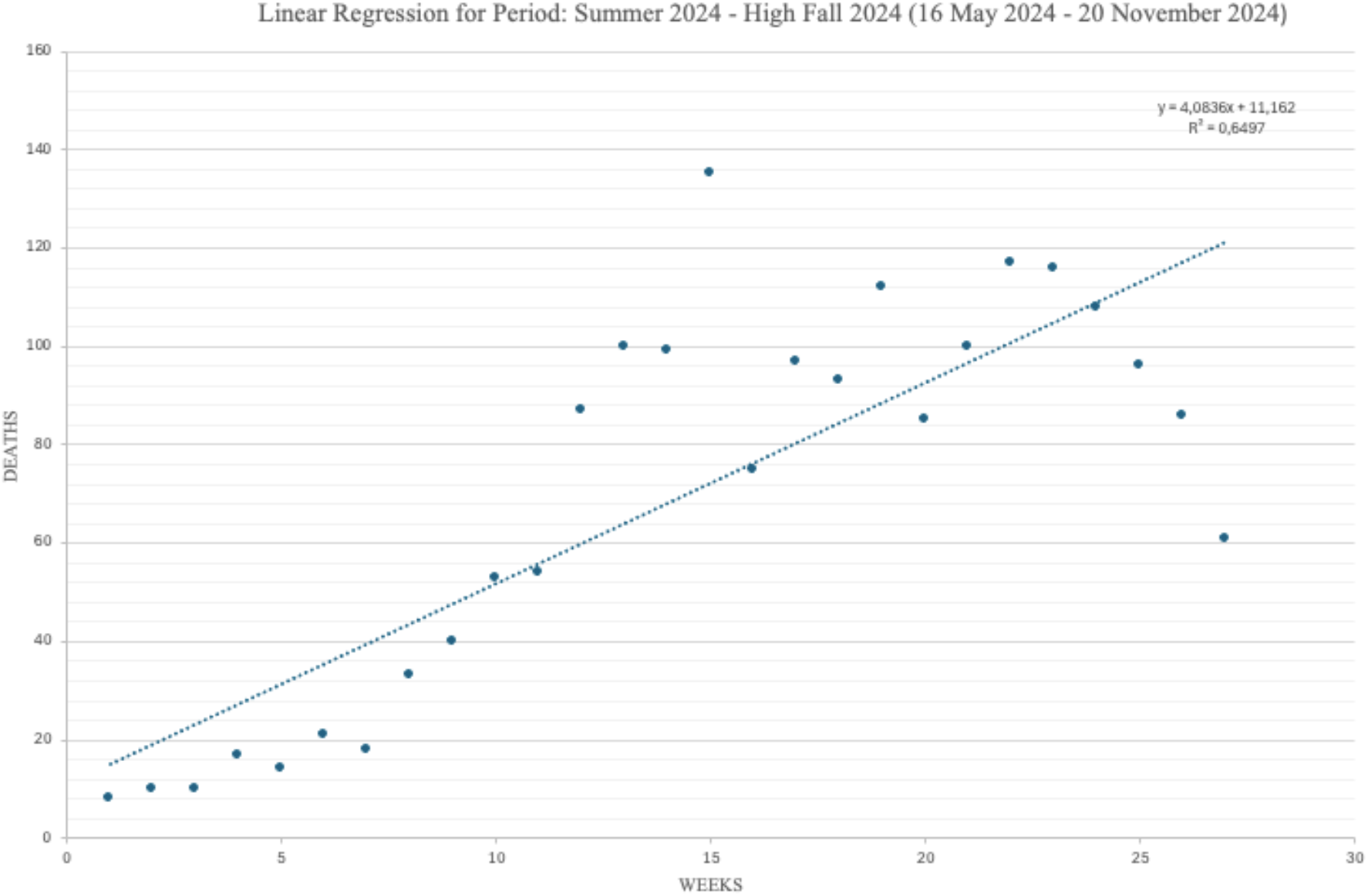
Period 1(Summer 2024/High Fall 2024): Regression segment. Slope coefficient *b* = +4.08. Coefficient of determination *r*^*2*^ = 64.97%

**Fig. 3.**
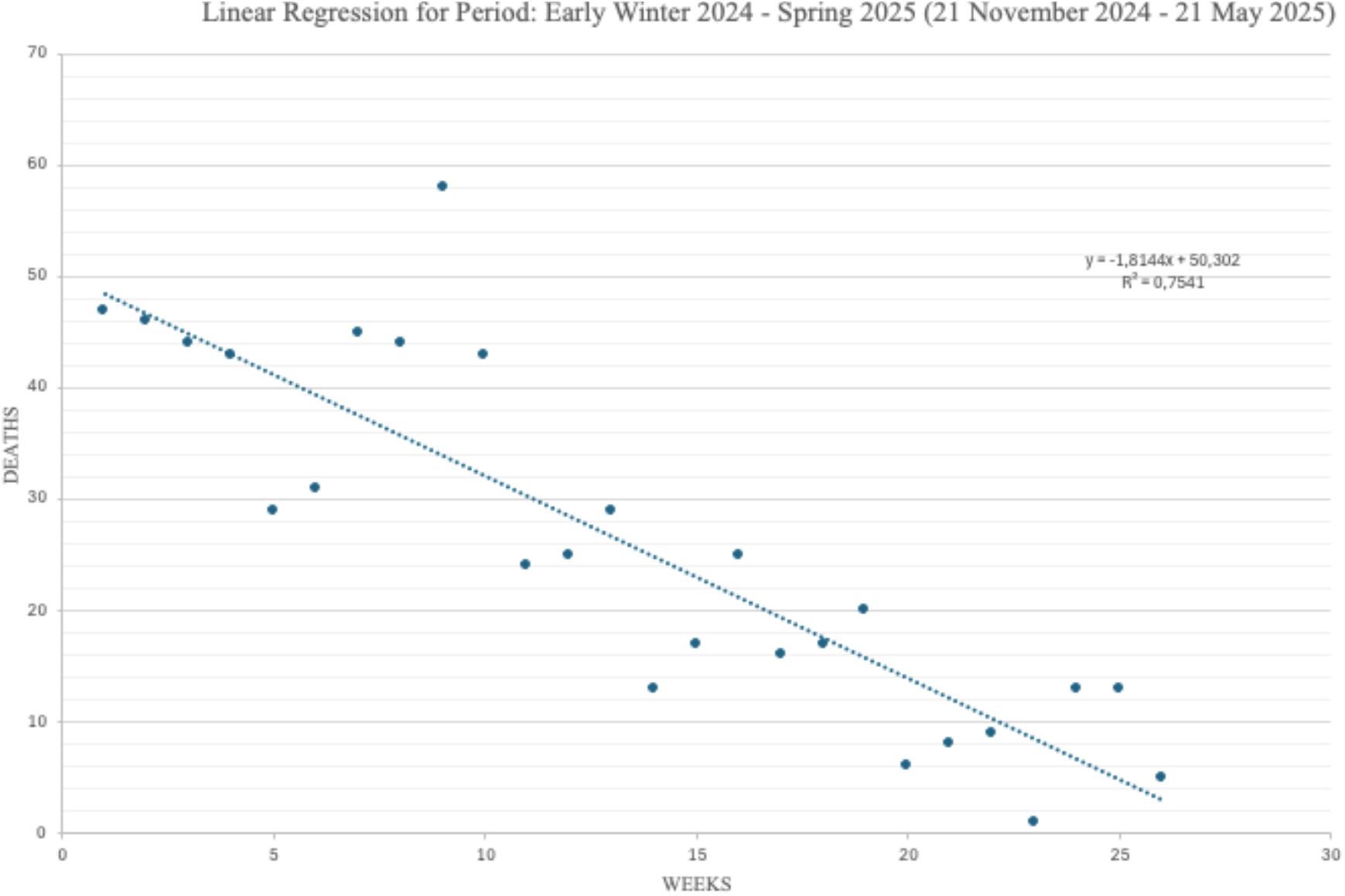
Period 2 (Early Winter 2024/Post Winter 2025): Regression segment. Slope coefficient *b* = −1.81. Coefficient of determination *r*^*2*^ = 75.41%.

In both the Figures, *Y* represents the number of weekly COVID-19 deaths for each given week (indicated over the *X* axis). Each Figure comes also with the values of the parameters *b* and *r*^*2*^, computed for each single segment and specified in the corresponding captions of the Figures. The *b* and *r*^*2*^ parameters of each period are also reported, for the sake of clarity, in Table 2, along with the average and the standard deviation values of the weekly COVID-19 deaths.

**Table 2.** Characteristics of the regression segments for the Periods of Figs. 2–3: Average and standard deviation of COVID-19 deaths per week, plus values of *b* and *r*^2^ per each period.

| Period | Season Type | Trend | Average<br>(Deaths/Week) | Std Deviation<br>(Deaths/Week) | Slope coefficient<br>$b$ | Determination<br>coefficient $r^2$ |
| --- | --- | --- | --- | --- | --- | --- |
| Period 1 | Summer –<br>High Fall | Increasing | 68.70 | 39.67 | +4.08 | 64.97% |
| Period 2 | Early Winter<br>– Spring | Decreasing | 25.80 | 15.67 | -1.81 | 75.41% |

As already anticipated, while the slope of a regression segment provides an immediate visual impression of the steepness of the corresponding COVID-19 mortality trend, the numerical value of *b* gives the exact number of COVID-19 deaths by which that trend increases/decreases with each new week. Finally, *r*^*2*^ informs on how well a given regression segment fits the available observations, in a scale from 0 to 1 (i.e., 0-100%).

Various factors are noteworthy by examining Figs. 2-3 and Table 2. First, COVID-19 deaths are more common in the period plotted in Fig. 1, with an increasing seasonal mortality pattern rising in late spring/early summer and peaking in high fall/early winter (and an average progressive increase of the number of deaths, per each new week, of almost 4). Second, a descending mortality trend is observable, instead, in the period portrayed in Fig. 2, starting to go down in early winter with a steady decrease until early spring (here, the average progressive reduction of the number of deaths per each new week is almost 2).

Final and most important is the fact that these seasonal mortality trends are clearly consistent throughout previous years (2021-2024), when similar seasonal mortality variations were observed in other studies [4, 9-12].

Concerning the issue of the goodness-of-fit of our regression model, it should be clear that we were not looking for the best-optimized solution, but for two regression segments able to guarantee an acceptable approximation of the available COVID-19 deaths data. In this sense, the values we got for the coefficients of determination *r*^*2*^, ranging in the interval [65-75%], can be considered a reasonable result, in the light of the fact that the specialized literature suggests as acceptable values *r*^*2*^ coefficients (equal to or) above the threshold of 60/65% [13].

In addition, it should be also considered that all the *p-values* obtained during the use of our model were below the statistical significance value of *α* = 0.05, thus providing a further confirmation of the validity of this analysis.

## 4. Conclusion

We have fitted a segmented linear regression model with the time series data of the COVID-19 deaths in the period May 2024 – May 2025. The aim was to document and statistically validate the seasonal variations and time trends of COVID-19 mortality in Italy. Our findings suggest the presence of an increasing mortality trend rising in late spring and early summer to peak in high fall/early winter. Then, we observe a steady decrease starting in mid-winter until almost the end of spring. These trends are in keeping with those observed in other studies [4], during the period 2021-2024, when the Omicron and post-Omicron variants were predominant. We recognize that we have avoided identifying the motivations behind these seasonal drifts that hence need further investigation [14-16]. We are aware that the method we used is not the one with which COVID-19 deaths are typically counted [17-24], but our target was just to look at how quickly COVID-19 mortality trends grew or declined. Our study permits a clear identification of the COVID-19 mortality trends for each season helping that subset of *at risk* population who can be seasonally vulnerable.

## Acknowledgements

Marco Roccetti expresses his gratitude to Eugenio De Rosa for the precious collaboration on earlier works.

